# Self-Configuring Capsule Networks for Brain Image Segmentation

**DOI:** 10.1101/2023.02.28.23286596

**Authors:** Arman Avesta, Sajid Hossain, Mariam Aboian, Harlan M. Krumholz, Sanjay Aneja

## Abstract

When an auto-segmentation model needs to be applied to a new segmentation task, multiple decisions should be made about the pre-processing steps and training hyperparameters. These decisions are cumbersome and require a high level of expertise. To remedy this problem, I developed self-configuring CapsNets (scCapsNets) that can scan the training data as well as the computational resources that are available, and then self-configure most of their design options. In this study, we developed a self-configuring capsule network that can configure its design options with minimal user input. We showed that our self-configuring capsule netwrok can segment brain tumor components, namely edema and enhancing core of brain tumors, with high accuracy. Out model outperforms UNet-based models in the absence of data augmentation, is faster to train, and is computationally more efficient compared to UNet-based models.

## Introduction

Despite the increasing popularity of deep-learning auto-segmentation methods, their implementation into clinical practice has been hindered by the need to configure their design options, which require a high level of expertise.^1,2^ If the design options of a deep-learning method are not optimally chosen for a particular task, the performance of the model drops significantly.^3^ This makes the process of adapting and training a deep-learning auto-segmentation model quite challenging, particularly in auto-segmenting 3D biomedical images where the image properties vary drastically.^3^ Examples of image properties that vary from one dataset to another include the image size, voxel spacing, voxel anisotropy, and the segmentation class ratios. An additional layer of complexity is posed by varying computational resources that are available for model training and deployment. Examples of these computational resources include the number of CPU cores, the amount of RAM, and the amount of GPU memory.

The properties of the input images and the computational resources that are available vary from one clinical setting to another, affecting the optimal design options that should be chosen for the training and deployment of an auto-segmentation model. Examples of the design options that should be chosen for each particular task, which depend on the input images and the computational resources that are available, include pre-processing steps such as image resampling and resizing strategy, patch size, class sampling strategy, batch size, and learning rate scheduling.^2^

Using empirical methods to choose the design options of deep-learning auto-segmentation models often leads to suboptimal design choices.^4^ Examples of the design options that should be chosen for each particular task, which depend on the input images and the computational resources that are available, include pre-processing steps such as image resampling and resizing strategy, data augmentation strategy, patch size, class sampling strategy, batch size, and learning rate scheduling strategy. Optimization methods proposed by previous studies in automatic machine learning (AutoML) are often not feasible due to the high-dimensional nature of the combinations of all design options.^4^ Therefore, these design options are often chosen by experts using an iterative trial-and-error process, which often leads to suboptimal auto-segmentation pipelines. Small errors in choosing the design options often leads to large drops in model performance.^1,2,4^

The problem of choosing the optimal design options for a model becomes even more cumbersome in the presence of real-world heterogeneous data. While most publicly available biomedical imaging datasets contain highly curated, high-resolution images with minimal imaging artifacts, the images in our clinical practice often have vastly different image resolution, image quality, varying degrees of voxel anisotropy, and imaging artifacts. In this setting, choosing the optimal design options for an auto-segmentation model would be difficult even for an experienced operator.^2,4^

In this study, I aim at automating the process of choosing these design options by developing a capsule network model that can scan the training data as well as the computational resources that are available, and then self-configure most of its design options. I propose that a self-configuring capsule network (scCapsNet) that does not need a human expert to optimize its design options would facilitate clinical implementation.

### The Yale Glioma Dataset

I used the images of 755 patients in the Yale Glioma Dataset that were scanned across several healthcare facilities within the Yale New Haven Health system. This dataset contains a wide variety of brain tumors (Table 5.1) that span benign grade 1 tumors to malignant grade 4 glioblastomas (Table 5.2). The images in this dataset are highly heterogeneous because they are scanned using 14 distinct MR scanners, dissimilar MR acquisition parameters, various scanning orientations (axial, sagittal, coronal, and 3D acquisitions), different voxel spacings, and various degrees of voxel anisotropy (Figure 5.1). I randomly split the patients in this dataset into training (603 MRI patients, 80% of data), validation (75 patients, 10% of data), and test (75 patients, 10% of data) sets. Table 5.2 provides patient demographics. This study was approved by the Institutional Review Board of Yale School of Medicine (IRB number 2000027592).

**Table 5.1:** Brain tumor types that are represented within the Yale Glioma Dataset.

| Diagnosis | Percentage |
| --- | --- |
| Glioblastoma | 50.6% |
| Anaplastic astrocytoma | 6.4% |
| Oligodendroglioma | 6.3% |
| Glioma, not otherwise specified | 4.1% |
| Pilocytic astrocytoma | 3.9% |
| Anaplastic oligodendroglioma | 2.4% |
| Astrocytoma | 2.2% |
| Diffuse astrocytoma | 1.7% |
| Oligoastrocytoma | 1.6% |
| Gliosarcoma | 0.3% |
| Pilomyxoid astrocytoma | 0.2% |
| Subependymal giant cell astrocytoma | 0.2% |
| Pleomorphic xanthoastrocytoma<br>0.2% |  |
| Low grade glial neoplasm | 0.1% |
| Tectal glioma | 0.1% |
| Ganglioglioma | 0.1% |
| Pilomyxoid astrocytoma WHO Grade 2 | 0.1% |
| Pylomyxoid astrocytoma | 0.1% |
| Desmoplastic infantile ganglioma/astrocytoma | 0.1% |
| Gliosarcoma | 0.1% |
| Anaplastic ependymal tumor | 0.1% |

**Table 5.2:**
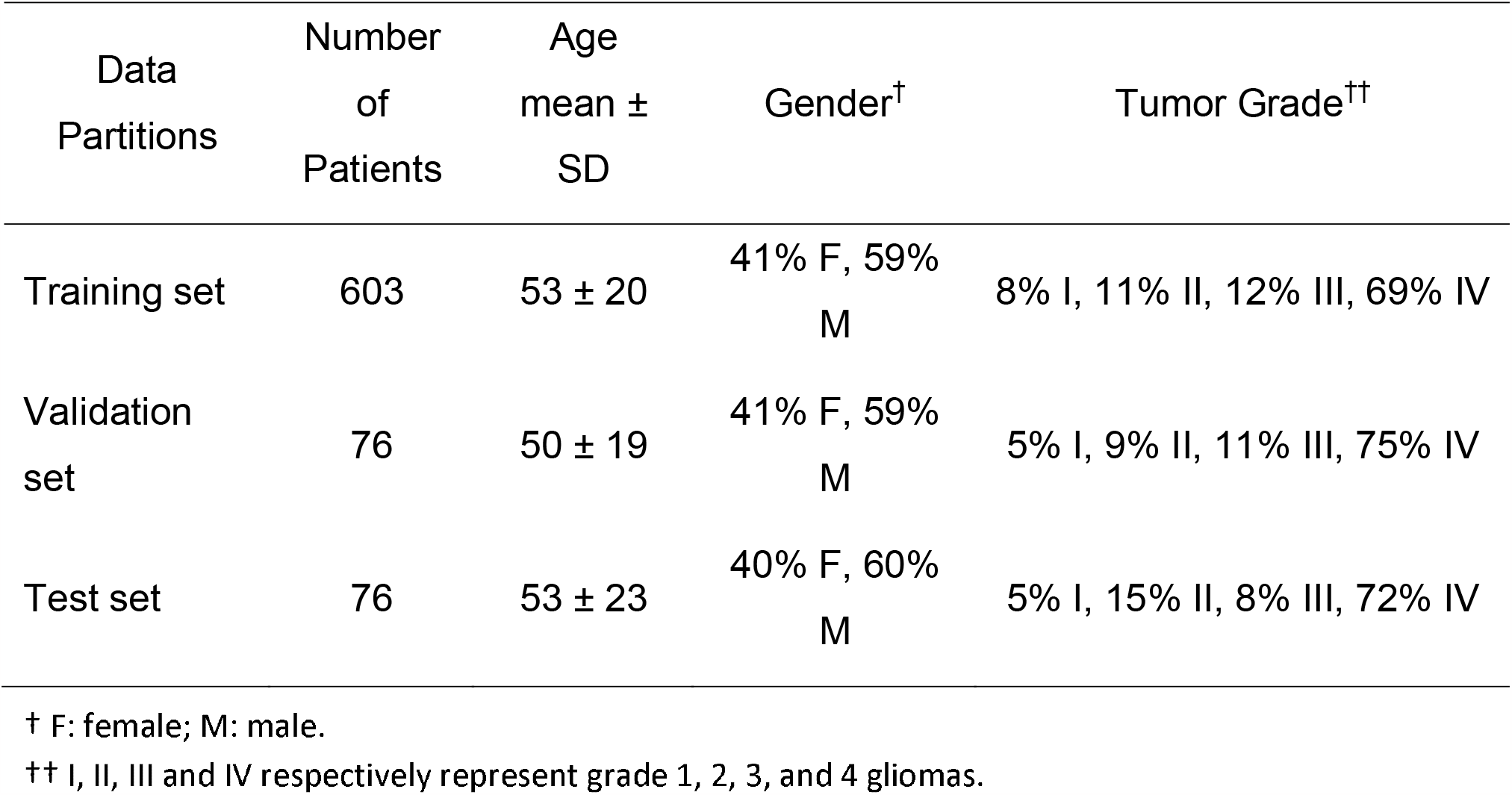
Study participants tabulated by the training, validation, and test sets.

**Figure 5.1:**
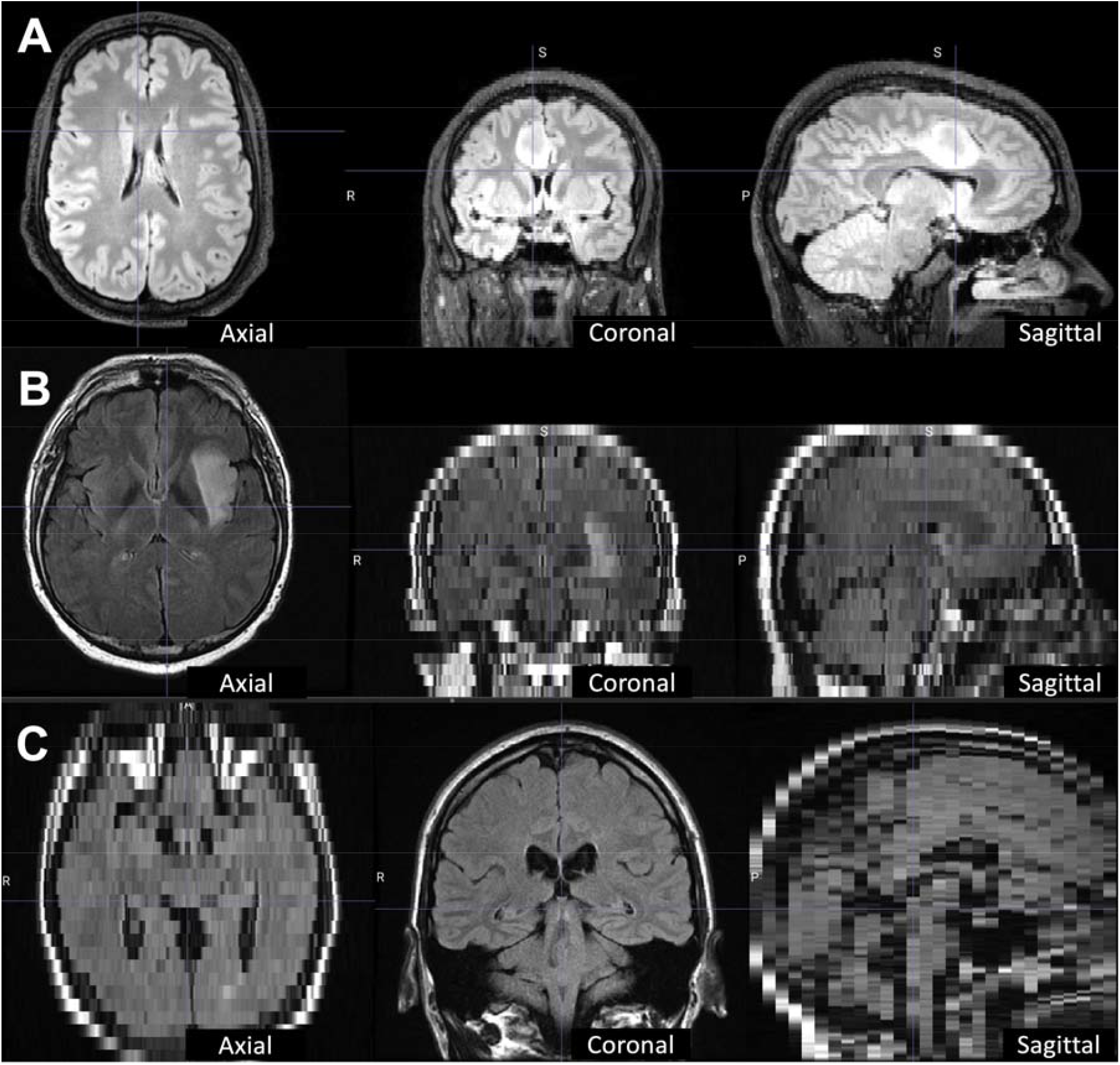
the Yale Glioma Dataset contains a large variety of MR images that were acquired using 16 MRI scanners with dissimilar MR acquisition parameters. For instance, the FLAIR images in this dataset include high-quality, high-resolution images with isotropic voxel spacing of 1×1×1 mm (A), axial MR acquisitions with high resolution in the axial plane but thick axial slices with voxel spacing of 0.4×0.4×7 mm (B), and coronal MR acquisitions with high resolution in the coronal plane but thick coronal slices with voxel spacing of 0.5×0.5×14 mm (C).

**Figure 5.2:**
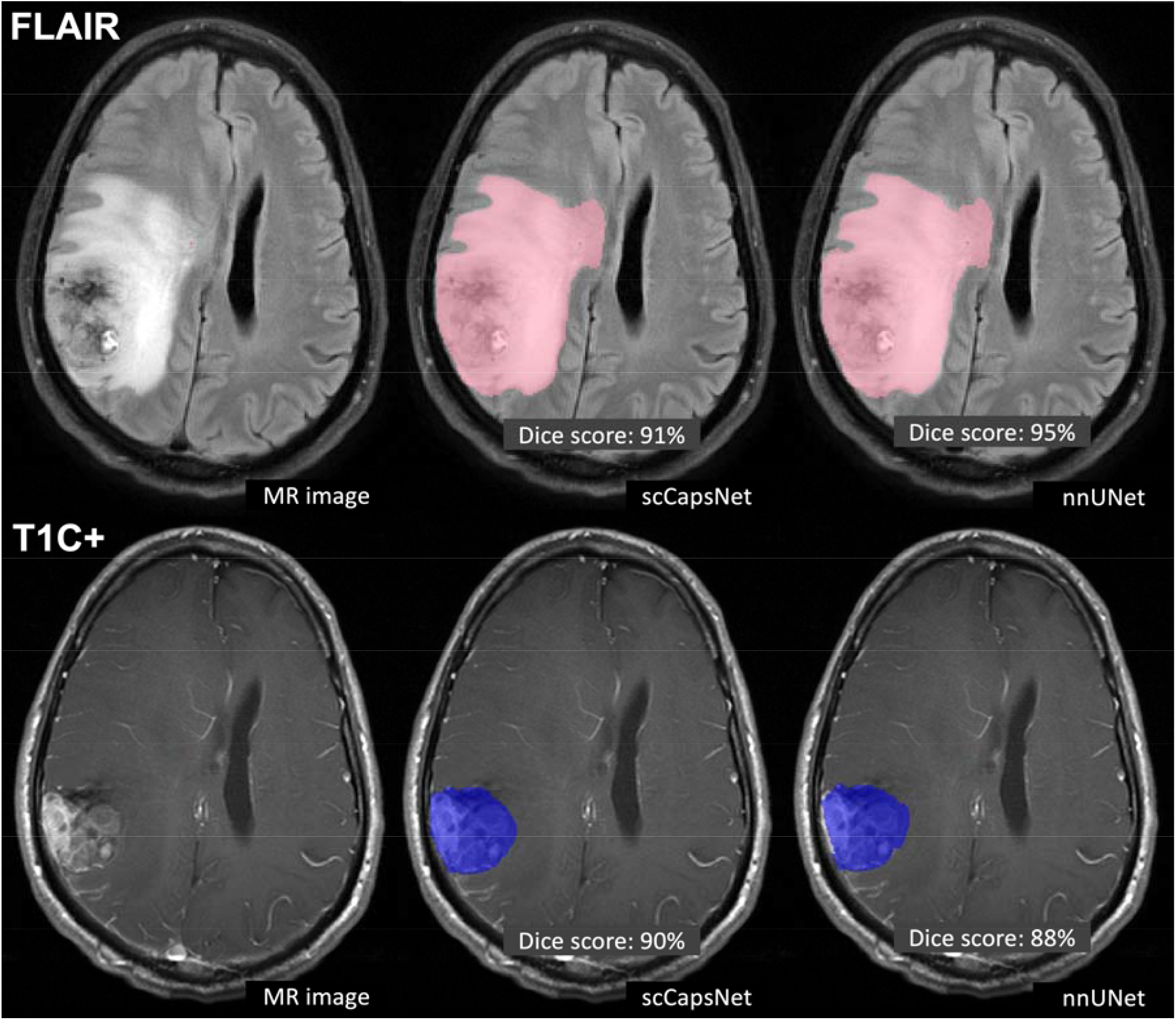
Comparing scCapsNet and nnUNet in auto-segmenting tumor components in a patient with glioblastoma. The top row shows auto-segmentation of the tumor edema/gliosis on the FLAIR image, and the bottom row shows auto-segmentation of the tumor enhancing core on the post-contrast T1-weighted (T1C+) image.

### Segmentation Targets: Components of Brain Tumors

I aimed at developing scCapsNet models that can segment two tumor components: 1) the tumor edema/gliosis on the fluid-attenuated inversion recovery (FLAIR) images; and 2) the tumor enhancing core on post-contrast T1-weighted images. Segmenting these two tumor components are clinically important for radiation therapy, neurosurgery, and treatment response monitoring.^5–7^

The tumors in this dataset were manually segmented within the picture archiving and communication system (PACS) of Yale New Haven Hospital. Five medical students, who were trained to manually segment the tumor components, manually segmented the images for 1,001 patients. Of these, the segmentations of 755 patients were checked, manually edited, and finalized by a board-certified attending neuroradiologist (Mariam S. Aboian, MD, PhD). I used this subgroup of 755 patients for training and testing my models.

### Automated Paradigm for Determining Computational Resources

The available computational resources determine key design options for image pre-processing and model training. For instance, a larger GPU memory allows for larger batch and patch sizes. To develop a self-configuring model, I used the following packages to automatically determine the computational resources that are available:

1. Number of CPU cores: I used the *multiprocessing* package in Python to read the number of available CPU cores.^8^ I set aside one CPU core for the operating system and used the remaining CPU cores for parallel image processing. Using this technique, I parallelized time-consuming pre-processing steps including bias field correction and image resampling, as well as time-consuming data loading steps during model training.
2. Available RAM: I used the *psutil* (process and system utilities) package in Python to read the amount of free RAM that is available for computing.^9^ To accelerate model training, I implemented a data loader that prepares a queue of input/output pairs using the CPU and stores this queue in RAM, ready to be used by the GPU to train the model. The length of this input/output queue is automatically determined by the amount of available RAM.
3. Available GPU memory: I used the NVIDIA Management Library’s *pynvml* package in Python to read the amount of free GPU memory that is available for computing.^10^ I defined configurations in my code that automatically increases the batch size and patch size if more GPU memory is available.

I have provided a sample code in Appendix 5 that shows how to use these three packages to read the available computational resources.

### Automated Paradigm for MRI Pre-Processing

I designed an automated pre-processing paradigm that can pre-process MRIs with different MR sequences, acquisition parameters, voxel spacings, and voxel anisotropies while requiring minimal user input. The steps of pre-processing include:

1. I used HD-BET for brain extraction, which a deep-learning method that can extract the brain from images that contain space-occupying lesions.^11^ To ensure that HD-BET correctly extracts the brain from images in the Yale Glioma Dataset, I randomly selected 100 MRIs and checked the brain-extracted images. All randomly-selected images showed correct brain extraction without major issues.
2. I used the Simple ITK package for bias field correction.^12^ Because bias field correction using this method is a time-consuming step, I parallelized this step by simultaneously processing multiple images using multiple CPU cores. The number of parallel processes is automatically determined by the model, depending on the available CPU cores and the amount of available RAM.
3. Each image and its accompanying ground-truth segmentation image are then cropped around the brain confines, thereby reducing their size.
4. The voxel intensities of the images are then normalized using Z-normalization.^13^
5. Label remapping is done if the segmentation labels are not encoded by consecutive numbers starting from 0. In label remapping, the segmentation labels are automatically remapped so that the background is encoded by 0 and the foreground labels are encoded by consecutive numbers starting from 1.^13^
6. Resampling is done to transform all images into the same voxel spacing. first, all images in the training set are scanned for their voxel spacing. Then, the median voxel spacing of all images is computed. Finally, all images in the dataset are resampled onto this median voxel spacing. For voxels that are isotropic or near-isotropic, third-order cubic interpolation is used for resampling. For voxels that are highly anisotropic, resampling in the direction that has the largest voxel spacing is done using linear interpolation (to prevent ghosting artifacts caused by thick-slice resampling). Segmentations are resampled using zero-order interpolation after one-hot-encoding. Zero-order interpolation for segmentation labels is necessary to prevent generation of non-integer numbers that do not represent any segmentation label.^13^
7. Finally, the quality of all pre-processing steps was checked for each image to ensure that the shape, voxel spacing, coordinate system, and the affine transform from the image space to the scanner space is consistent between each raw image and its accompanying ground-truth segmentation image. Finally, the preprocessed images and segmentation masks are saved as NIfTI files.^14^

### Accelerated Data Loading

To make the model training faster, I implemented a data loading method that loads the input image and the accompanying segmentation, randomly samples patches from them, forms data batches, and finally forms a queue of input/output batches that are ready to be used by the GPU for model training. These computations are done using the CPU and RAM while the model is trained on the GPU in parallel. Because the CPU and RAM prepare input/output pairs at the same time that the GPU trains the model, the two processes do not wait for each other, resulting in accelerated training that is about twice faster.^13^ Additionally, several CPU cores are recruited to prepare the input/output pairs in parallel. The length of the inputs/outputs queue is automatically choses depending on the batch size, patch size, and the amount of available RAM.

### Comparing Self-Configuring Capsule Networks with nnUNets and UNets

In addition to training the self-configuring capsule network, I also trained nnUNet and UNet models using the same training data,^15–18^ followed by comparing their segmentation accuracy and computational efficiency using the same test data. Because the nnUNet has its own pre-processing pipeline, I fed the nnUNet with raw images and accompanying segmentation.

Because I wanted to compare the performance of scCapsNet with UNet-based models in the absence of data augmentation and because the nnUNet does data augmentation automatically, I also trained and tested a UNet using the data that was pre-processed by the scCapsNet pre-processing pipeline. The architecture and training hyperparameters of the UNet are provided in Chapter 3 and Appendix 3.

### Auto-Segmentation Performance Metrics

I compared the segmentation accuracy of the scCapsNet with nnUNet in auto-segmenting the tumor edema/gliosis on FLAIR images as well as the tumor enhancing core on post-contrast T1-weighted images. Segmentation accuracy was quantified using Dice scores. To compare the segmentation accuracy of scCapsNet with UNet-based models in the absence of data augmentation, I trained and tested a UNet using the pre-processed data that was used to train the scCapsNet without data augmentation. To compare the computational speed during training, I compared the time that the scCapsNet and nnUNet would need to converge to the Dice score of 80% over the validation set. To compare the computational speed during deployment, I compared the time that the scCapsNet and nnUNet would need to pre-process and segment a brain MRI. Finally, I compared the GPU memory that is required by the scCapsNet and nnUNet models.

### Software and Hardware used for Model Implementation

Image pre-processing was done using Python (version 3.10), SimpleITK (version 2.2.0),^12^ TorchIO (version 0.18.78),^13^ NiBabel (version 5.0.0),^19^ and HD-BET.^11^ PyTorch (version 1.13.1) was used for model development and testing. SciPy (version 1.6.0) was used for statistical testing. Training and testing of the models were run on GPU-equipped servers (4 vCPUs, 16 GB RAM, 16 GB NVIDIA GPU). The code used to train and test our models is available on our lab’s GitHub page:

https://github.com/Aneja-Lab-Yale/Aneja-Lab-Public-scCapsNet.

### Auto-Segmentation Performance Results

The nnUNet slightly outperformed the scCapsNet model in auto-segmented the tumor edema/gliosis on FLAIR images, with average Dice scores of 86% and 89% for the two models, respectively. The scCapsNet slightly outperformed the nnUNet in auto-segmenting the tumor enhancing core on post-contrast T1-weighted images, with average Dice scores of 89% and 88%, respectively. However, none of the differences between the two models were statistically significant (Table 5.3).

**Table 5.3:**
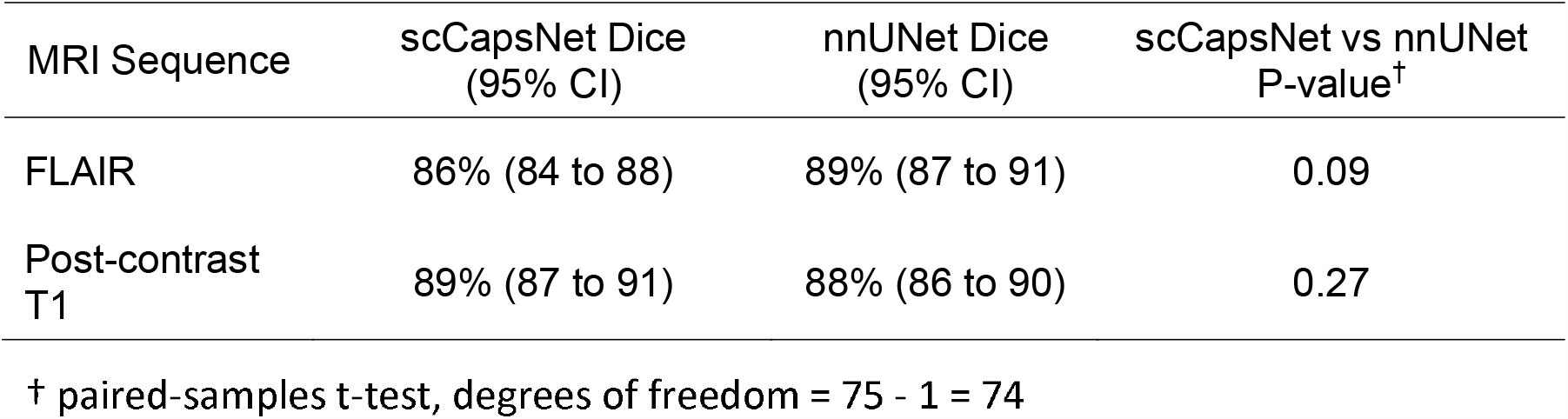
Comparing the performance of scCapsNet and nnUNet in segmenting tumor edema/gliosis on FLAIR images, and the enhancing core of the tumor on post-contrast T1-weighted images.

In the absence of data augmentation, there was a drop in the performance of the UNet-based models. The UNet auto-segmented the FLAIR and post-contrast T1-weighted images with Dice scores of 84% and 85%, respectively. The scCapsNet outperformed the UNet-based models in the absence of data augmentation (Table 5.4).

**Table 5.4:**
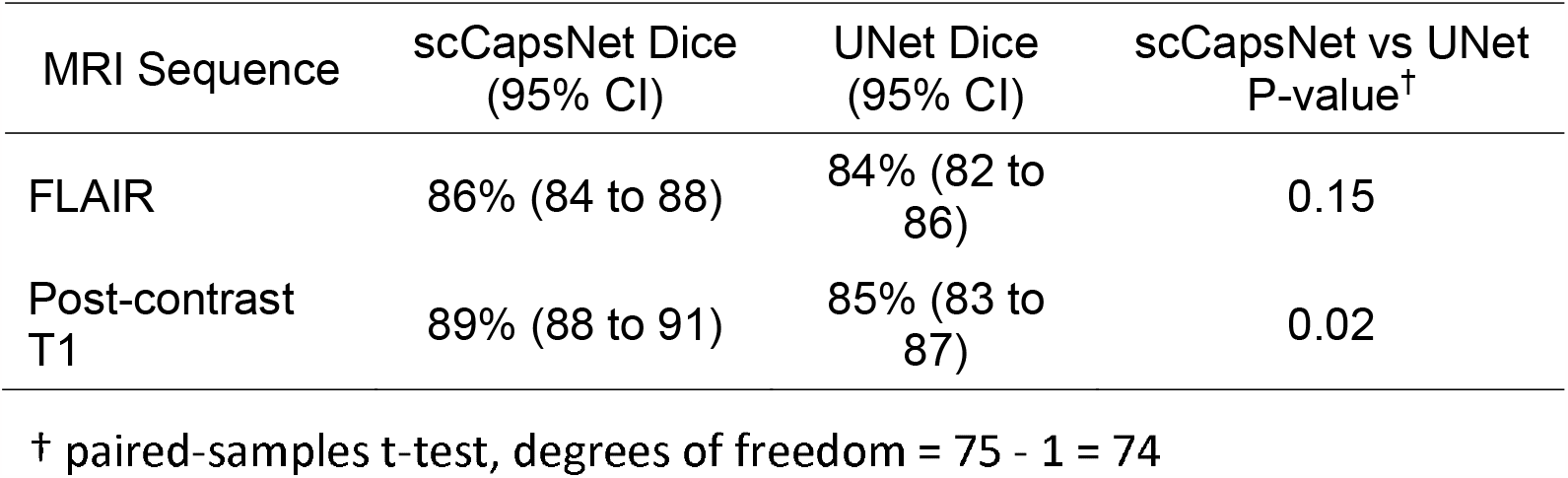
Comparing the performance of scCapsNet and UNet without data augmentation.

The scCapsNet converges faster during training compared to nnUNet. During training, the scCapsNet and nnUNet models reached the Dice score of 80% over the validation set after 13 and 38 hours, respectively (Figure 5.3). During deployment, the scCapsNet and nnUNet respectively require 4 and 3 minutes to pre-process and segment a brain MRI. Out of the 4 minutes, the scCapsNet spends more than 3 minutes on two pre-processing steps: brain extraction and bias field correction.

**Figure 5.3:**
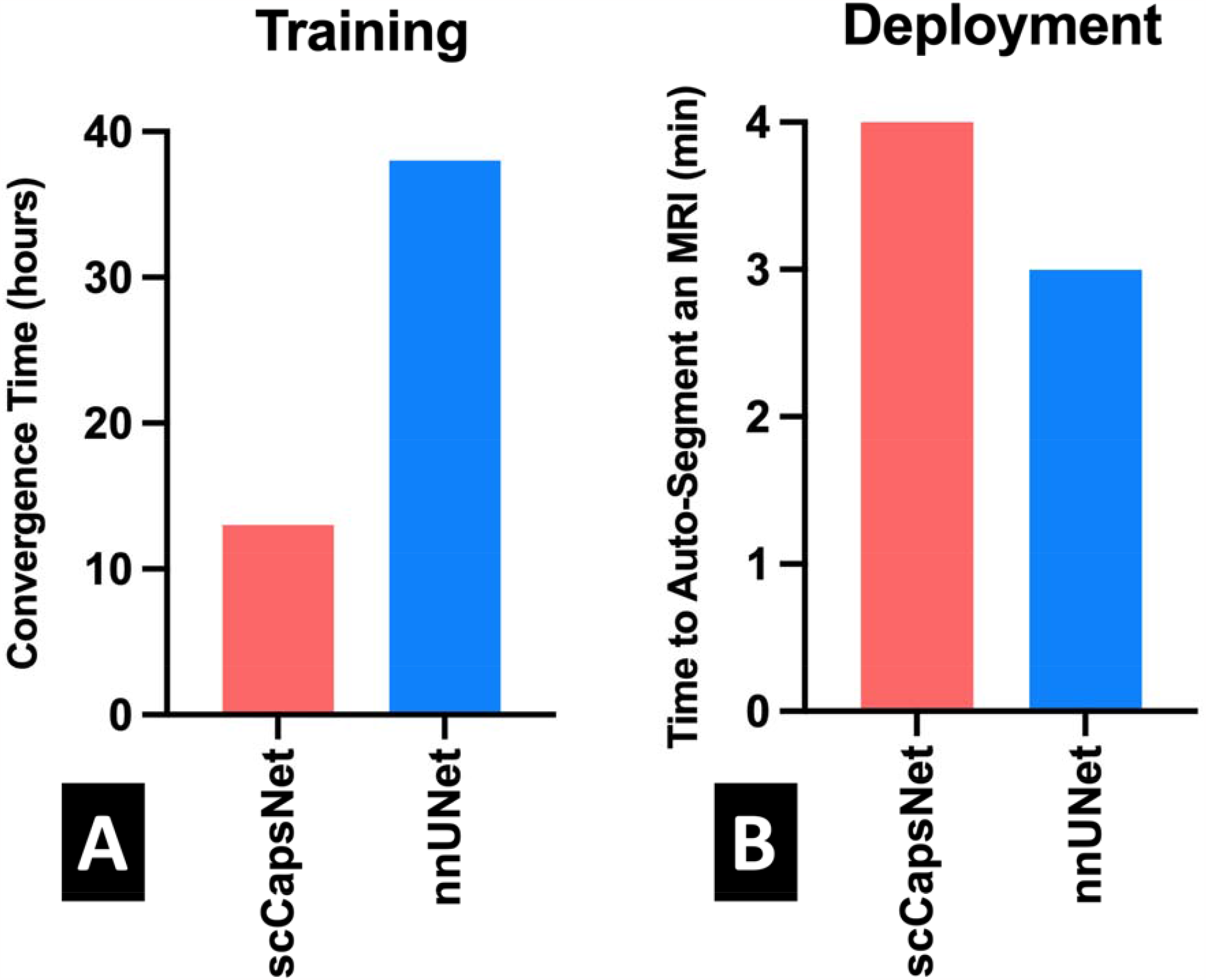
Comparing the computational speed between the scCapsNet and nnUNet models. The scCapsNet converges faster during training and reaches the Dice score of 80% after 13 hours, while the nnUNet reaches this Dice score after 38 hours. During deployment, the scCapsNet and nnUNet respectively require 4 and 3 minutes to pre-process and segment a brain MRI. The scCapsNet is slower during deployment because two pre-processing steps, namely skull stripping and bias field correction, are slow processes that take more than 3 minutes to complete. Notably, the nnUNet does not perform these two pre-processing steps.

The scCapsNet is computationally more efficient compared to nnUNet. The scCapsNet and nnUNet respectively have 7,400 and 90,300 trainable parameters, which respectively occupy 28 and 345 megabytes on the GPU memory. The total sizes of the scCapsNet and nnUNet are respectively 5 and 31 gigabytes (Figure 5.4).

**Figure 5.4:**
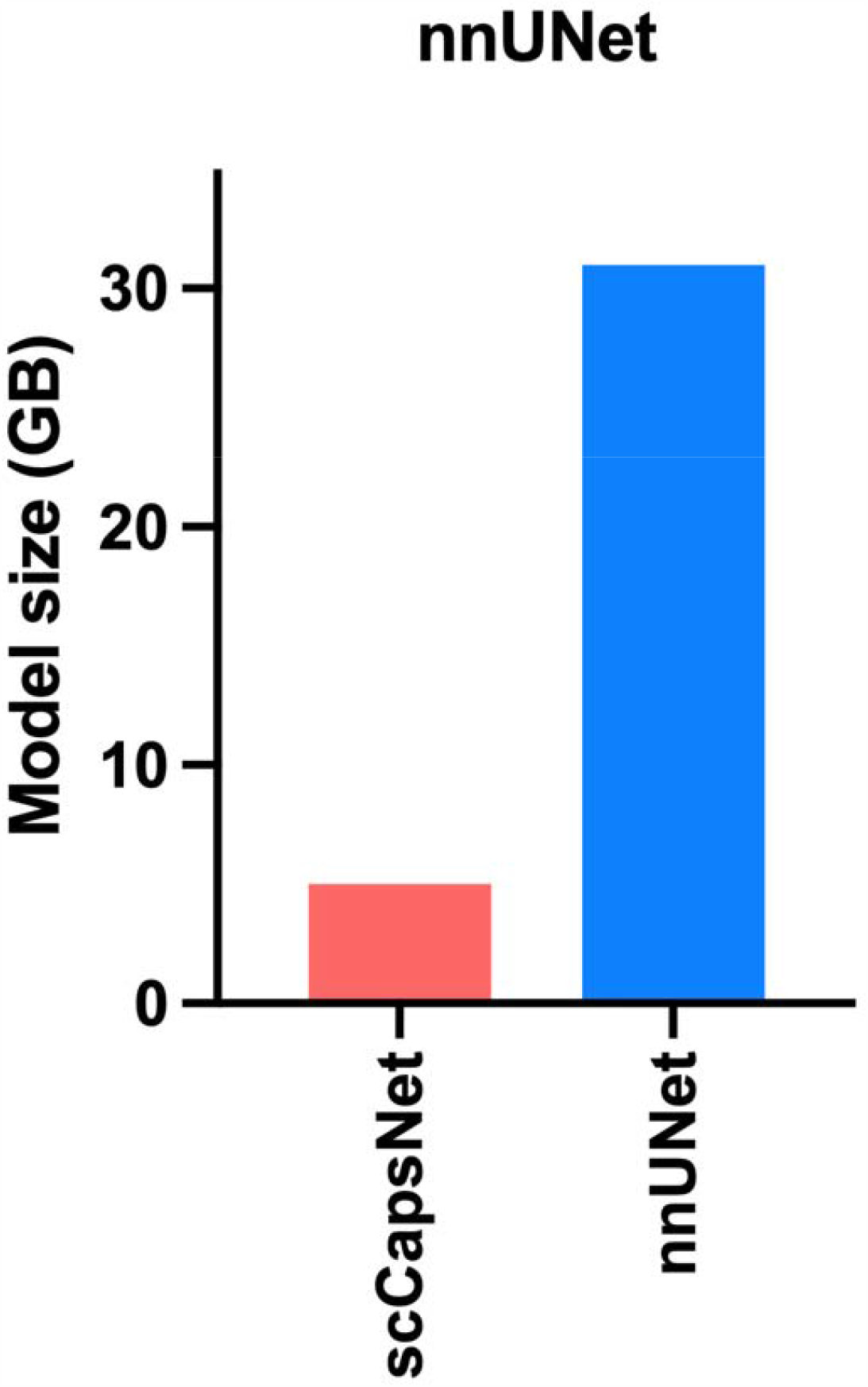
Comparing the GPU memory required by the scCapsNet and nnUNet models. The bars represent the computational memory required to accommodate the total size of each model, including the parameters plus the cumulative size of the forward- and backward-pass feature volumes. The total sizes of the scCapsNet and nnUNet models are respectively 5 and 31 gigabytes.

### Brain MRI Auto-Segmentation using Self-Configuring Capsule Networks

In this chapter, I aimed at developing a self-configuring capsule network (scCapsNet) that is feasible for clinical implementation. The scCapsNet can self-configure its design options by considering the images that should be auto-segmented as well as the computational resources that are available. I showed that the scCapsNet can segment brain tumor components with high accuracy. Additionally, I showed that the scCapsNet outperforms UNet-based models in the absence of data augmentation, is faster to train, and is computationally more efficient compared to UNet-based models.

My results extend the prior literature in key ways. I developed the first self-configuring capsule network and comprehensively benchmarked its segmentation accuracy and computational efficiency. To train and test my models, I used a large dataset of clinical MRIs that more closely represent real-life MRIs in our clinical practice, with the presence of suboptimal image qualities and MRI artifacts. Using this dataset, I compared the performance of the scCapsNet against the most successful auto-segmentation model that is currently used, namely nnUNet.^2^ Therefore, my results provide practical benchmarking between the two methods on real-life clinical MRI.

My results corroborate previous studies showing that self-configuring auto-segmentation models are highly valuable for clinical implementation.^1,2,20–29^ Isensee et al developed the nnUNet model, which is a self-configuring model based on UNets.^2^ While the nnUNet also self-configures its design options by considering the data and the computational resources, the processes by which the nnUNet chooses its design options are different from my approach. The nnUNet model runs a few experiments at the start of the training, changing the patch size, batch size, and model depth until the GPU memory runs out. These experiments determine the largest patch size, batch size, and model depth that the GPU memory can accommodate.

The scCapsNet, unlike the nnUNet, directly reads the computational resources through Python modules, without the need to run any experiments. The computational resources that are read are the GPU memory, RAM capacity, and the number of CPU cores. Then, the model assigns the appropriate patch size, batch size, length of the inputs/outputs queue that are prepared by the CPU during training, and the number of parallel CPU cores that are recruited, given the computational resources that are available. Notably, the nnUNet model requires user input to determine the number of CPU cores that should be recruited. I propose that my approach is computationally more efficient. Additionally, the nnUNet does not perform skull stripping and bias field correction during pre-processing.^2^

The scCapsNet converges faster during training because it has fewer trainable parameters and because of the accelerated data loading methods that are implemented into the scCapsNet model. The accelerated data loading paradigm decreases the training time by a factor of two. However, the scCapsNet is slightly slower than the nnUNet during deployment, since it requires four minutes to pre-process and segment a brain MRI. Of these four minutes, more than three minutes are spent on two pre-processing steps: skull stripping and bias field correction. Notably, the nnUNet model does not perform these two steps during pre-processing. I am currently studying the necessity of brain extraction and bias field correction during pre-processing. As I will discuss in Chapter 6, I am also studying the use of data augmentation techniques to simulate bias field inhomogeneities so that we can feed the scCapsNet model with images that contain bias field inhomogeneities without the need for correcting them.

My experiments have several notable limitations. First, I only compared the performance of scCapsNet and nnUNet models in segmenting brain tumors on MR images. The results of my experiments may not generalize to other brain pathologies, imaging modalities, or other body organs. Second, I used the Yale Glioma Dataset benchmark the performance of my models. My results may not generalize to other healthcare facilities with different patient populations, dissimilar distributions of brain tumors, or different MR scanners. Nonetheless, the images in the Yale Glioma Dataset represent a wide range of brain gliomas, MR scanners, and MR acquisition parameters. Finally, the comparisons between the computational speed of scCapsNet and nnUNet models depend on the computational resources that are available. These comparisons may not generalize to other computational settings. However, I used computational resources that are commonplace in the deep learning computing units.

## Conclusion

In this chapter, I developed a self-configuring capsule network that can auto-segment brain MRIs with minimal, if any, user input. I showed that this model can segment the components of brain tumors with high accuracy and is computationally more efficient that currently used auto-segmentation models. In the next chapter, I will explore how this model, or any auto-segmentation model, can be brought to the bedside to help our patients.

## Data Availability

All data produced in the present study are available upon reasonable request to the authors.

## References

1. Heidenreich JF, Gassenmaier T, Ankenbrand MJ, et al. Self-configuring nnU-net pipeline enables fully automatic infarct segmentation in late enhancement MRI after myocardial infarction. Eur J Radiol 2021;141:109817.

2. Isensee F, Jaeger PF, Kohl SAA, et al. nnU-Net: a self-configuring method for deep learning-based biomedical image segmentation. Nat Methods 2021;18:203–11.

3. Litjens G, Kooi T, Bejnordi BE, et al. A survey on deep learning in medical image analysis. Med Image Anal 2017;42:60–88.

4. Hutter F, Kotthoff L, Vanschoren J, eds. Automated Machine Learning: Methods, Systems, Challenges. Cham: Springer International Publishing; 2019.

5. Mendel JT, Jaster AW, Yu FF, et al. Fundamentals of Radiation Oncology for Neurologic Imaging. RadioGraphics 2020;40:827–58.

6. Dundar TT, Yurtsever I, Pehlivanoglu MK, et al. Machine Learning-Based Surgical Planning for Neurosurgery: Artificial Intelligent Approaches to the Cranium. Front Surg 2022;9:863633.

7. Bennett EE, Angelov L, Vogelbaum MA, et al. The Prognostic Role of Tumor Volume in the Outcome of Patients with Single Brain Metastasis After Stereotactic Radiosurgery. World Neurosurg 2017;104:229–38.

8. Multiprocessing: process-based parallelism. URL: https://docs.python.org/3/library/multiprocessing.html. Accessed 2023-02-15. Python Doc.

9. Rodola G. Psutil: a cross-platform library for process and system monitoring in Python. URL: https://github.com/giampaolo/psutil. Accessed 2023-02-15.

10. Pynvml: Python Bindings for the NVIDIA Management Library. URL: https://pypi.org/project/pynvml. Accessed 2023-02-15.

11. Isensee F, Schell M, Pflueger I, et al. Automated brain extraction of multisequence MRI using artificial neural networks. Hum Brain Mapp 2019;40:4952–64.

12. N4 Bias Field Correction — SimpleITK 2.0rc2 documentation. URL: https://simpleitk.readthedocs.io/en/master/link_N4BiasFieldCorrection_docs.html. Accessed 2023-02-15.

13. Pérez-García F, Sparks R, Ourselin S. TorchIO: A Python library for efficient loading, preprocessing, augmentation and patch-based sampling of medical images in deep learning. Comput Methods Programs Biomed 2021;208:106236.

14. Li X, Morgan PS, Ashburner J, et al. The first step for neuroimaging data analysis: DICOM to NIfTI conversion. J Neurosci Methods 2016;264:47–56.

15. Cardenas CE, Yang J, Anderson BM, et al. Advances in Auto-Segmentation. Semin Radiat Oncol 2019;29:185–97.

16. Rudie JD, Weiss DA, Colby JB, et al. Three-dimensional U-Net Convolutional Neural Network for Detection and Segmentation of Intracranial Metastases. Radiol Artif Intell 2021;3:e200204.

17. Rauschecker AM, Gleason TJ, Nedelec P, et al. Interinstitutional Portability of a Deep Learning Brain MRI Lesion Segmentation Algorithm. Radiol Artif Intell 2022;4:e200152.

18. Weiss DA, Saluja R, Xie L, et al. Automated multiclass tissue segmentation of clinical brain MRIs with lesions. NeuroImage Clin 2021;31:102769.

19. NiBabel: Neuroimaging in Python. URL: https://nipy.org/nibabel/. Accessed 2023-02-16.

20. Alqaoud M, Plemmons J, Feliberti E, et al. nnUNet-based Multi-modality Breast MRI Segmentation and Tissue-Delineating Phantom for Robotic Tumor Surgery Planning. Annu Int Conf IEEE Eng Med Biol Soc IEEE Eng Med Biol Soc Annu Int Conf 2022;2022:3495–501.

21. Pettit RW, Marlatt BB, Corr SJ, et al. nnU-Net Deep Learning Method for Segmenting Parenchyma and Determining Liver Volume From Computed Tomography Images. Ann Surg Open Perspect Surg Hist Educ Clin Approaches 2022;3:e155.

22. Zhu Y, Chen L, Lu W, et al. The application of the nnU-Net-based automatic segmentation model in assisting carotid artery stenosis and carotid atherosclerotic plaque evaluation. Front Physiol 2022;13:1057800.

23. Li F, Sun L, Lam K-Y, et al. Segmentation of human aorta using 3D nnU-net-oriented deep learning. Rev Sci Instrum 2022;93:114103.

24. El-Hariri H, Souto Maior Neto LA, Cimflova P, et al. Evaluating nnU-Net for early ischemic change segmentation on non-contrast computed tomography in patients with Acute Ischemic Stroke. Comput Biol Med 2022;141:105033.

25. Ferrante M, Rinaldi L, Botta F, et al. Application of nnU-Net for Automatic Segmentation of Lung Lesions on CT Images and Its Implication for Radiomic Models. J Clin Med 2022;11:7334.

26. Zhu H, Yu H, Zhang F, et al. Automatic segmentation and detection of ectopic eruption of first permanent molars on panoramic radiographs based on nnU-Net. Int J Paediatr Dent 2022;32:785–92.

27. Zhang J, Li Z, Yan S, et al. An Algorithm for Automatic Rib Fracture Recognition Combined with nnU-Net and DenseNet. Evid-Based Complement Altern Med ECAM 2022;2022:5841451.

28. Dot G, Schouman T, Dubois G, et al. Fully automatic segmentation of craniomaxillofacial CT scans for computer-assisted orthognathic surgery planning using the nnU-Net framework. Eur Radiol 2022;32:3639–48.

29. Huo L, Hu X, Xiao Q, et al. Segmentation of whole breast and fibroglandular tissue using nnU-Net in dynamic contrast enhanced MR images. Magn Reson Imaging 2021;82:31–41.

